# Diminishing Marginal Benefit of Social Distancing in Balancing COVID-19 Medical Demand-to-Supply

**DOI:** 10.1101/2020.04.09.20059550

**Authors:** Pai Liu, Payton Beeler, Rajan K. Chakrabarty

## Abstract

Social distancing has been adopted as a non-pharmaceutical intervention to prevent the COVID-19 pandemic from overwhelming the medical resources across the United States (US). The catastrophic socio-economic impacts of this intervention could outweigh its benefits if the timing and duration of implementation are left uncontrolled and ill-strategized. Here we investigate the dynamics of social distancing on age-stratified US population and benchmark its effectiveness in reducing the burden on hospital and ICU beds. Our findings highlight the diminishing marginal benefit of social distancing, characterized by a linear decrease in medical demands against an exponentially increasing social distancing duration. We determine an optimal intermittent social-to-no-distancing ratio of 5:1 corresponding to ∼80% reduction in healthcare demands – beyond this ratio, benefit of social distancing diminishes to a negligible level.

**COVID-19 Medical Demand Forecast:** https://eece.wustl.edu/chakrabarty-group/covid/

## Introduction

In December 2019, a novel coronavirus named SARS-CoV-2 began infecting residents of Wuhan, China (*1-3*). SARS-CoV-2 causes moderate to severe respiratory symptoms that can progress to severe pneumonia (coronavirus disease 2019, COVID-19) (*4*). Despite the extreme disease containment measures taken in China (*5*), COVID-19 has spread rapidly to numerous countries and evolved into a global pandemic (*1, 3*). On January 30, 2020, the World Health Organization declared a “public health emergency of international concern” (*6*), and on the following day the United States Department of Health and Human Services declared a public health state of emergency (*7*).

During the week of February 23, the US Centers for Disease Control (US-CDC) reported new confirmed cases of COVID-19 in California, Oregon, and Washington, indicating the onset of “community spread” across the US (*7*). Until March 2, the total number of confirmed active COVID-19 cases in the US were 33, with new cases emerging in states of Texas, Arizona, Wisconsin, Illinois, Florida, New York, Rhode Island, and Massachusetts (*8*). In the following two weeks, this number has rapidly increased to 527 confirmed cases on March 9, and then to 4,216 cases on March 16 (*8*). State of California and New York have respectively declared state emergency on March 4 and 7 (*9, 10*). The White House declared national emergency on March 13 (*11*). Thus, major outbreak of COVID-19 epidemic across the US is inevitable. As of March 27, the total number of confirmed cases in the US has exceeded 100,000, surpassing that in China and Italy (*8*). The US-CDC asserted that the progression of COVID-19 in the US is still in its acceleration phase, with the epidemic peak yet to arrive (*7*). To prevent the rapid disease spread and meet the medical demands, social distancing has been adopted as a non-pharmaceutical measure across the country. Such a non-pharmaceutical intervention aims to slows down epidemic progression, and ultimately prevents the country’s medical system from collapsing due to overburdening of COVID-19 patients. However, its effectiveness and cost benefits as a function of implementation duration, timing, and strategy, especially in the context of COVID-19 epidemic, remain uncertain. Here, we comprehensively investigate the effectiveness of various social distancing practices, as well as their implementation strategies, on the reduction in the peak demand of medical resources, such as hospital beds and ICU beds (*12*).

## Methods

Our metapopulation epidemiological model involves the susceptible, exposed, infected, and recovered (SEIR) dynamics (*1,2,13,14*), the age-stratified disease transmissibility (*15,16*), and the possible largescale undocumented transmission (*17*) taking place in the 50 US states, Washington DC, and Puerto Rico (hereafter, they are generically denoted as states). For each state *n*, the local population was classified into four categories – susceptible, exposed, infected, and recovered – with the fraction of population within each category denoted as *s*_*n*_, *e*_*n*_, *j*_*n*_, and *r*_*n*_, respectively (*18*). The infected category was further divided into two sub-classes: the infected-and-documented 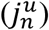 and the infected-and-undocumented 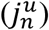, i.e. 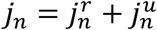 This treatment accounts for the substantial influence of the asymptomatic (or mildly symptomatic) COVID-19 carriers on accelerating the epidemic spread (*17*). Furthermore, the population within each category of *s*_*n*_, *e*_*n*_, 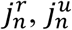 and *r*_*n*_ were divided into nine age-stratified compartments (hereafter age groups), in light of the strong age-dependent hospitalization rate of the COVID-19 patients (*12*). Specifically, age group *i* (with 1 ≤ *i* ≤ 8) comprised of individuals aged between 10*(*i*-1) and 10\**i*-1 years, while the 9^th^ age group included everyone aged 80 years and above. This age-stratification can be expressed as: *u*_*n*_ = ∑_*i*_ *u*_*n,i*_ (here, *u* is used to generically denote *s, e, j*^*r*^, *j*^*u*^, and *r*). Finally, the interstate exchange of individual within the *s*_*n*_, *e*_*n*_, 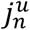, and *r*_*n*_ category was captured using a mobility matrix of *P*_*m,n*_ quantifying the probability that an individual leaving state *n* ends up in *m* (*18-20*).

The governing equations of our model can therefore be expressed as a set of first-order differential equations with respect to time (*tt*):

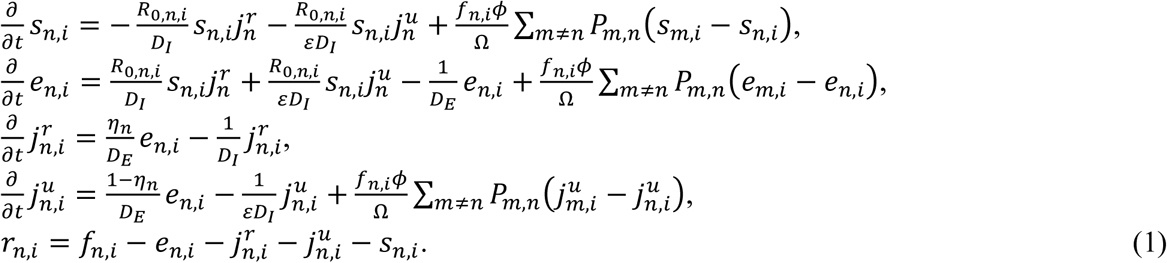

Here, *R*_0,*n,i*_ is an age-specific reproduction ratio representing the efficiency by which the COVID-19 (within state *n*) is transmitted to local population in age group *i*. The values of *R*_0,*n,i*_ were mapped from a state-wise reproduction ratio (*R*_0,*n*_) per the assumption that the disease transmissibility between age groups *i* and *k* is directly proportional to the average daily time-of-exposure (*T*_*i,k*_) amongst their members: *R*_0,*n,i*_ = *R*_0,*n*_ ∑_*k*_ *T*_*i,k*_/*q*, where *q* is a proportionality factor taking values of the largest eigenvalue of the *T*_*i,k*_ matrix (*15,16,21*). *D*_*E*_ and *D*_*I*_ respectively represent the mean incubation period and mean infectious period of COVID-19 (*1, 2, 22*); *η*_*n*_ represents local documentation ratio of the infected individuals, and ε is a constant factor denoting the mean elongation of infectious period for those undocumented COVID-19 carriers (*17*); *f*_*n,i*_ is the fraction of local population within age group *i, ϕ* is the daily passenger flux of the entire traffic network, and Ω represents the total US population (*18, 23*). The ratio *f*_*n,i*_ *ϕ*/Ω can be regarded as an interstate mobility parameter for members of age group *i*.

Our model was initialized on March 19 (*t*_0_) with the documented active COVID-19 cases 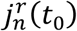 acquired from a web-based dashboard for real-time epidemic tracking published by Johns Hopkins University (*8*). Next, we estimated the initial number of undocumented cases, per 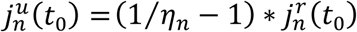, and the exposed cases, per 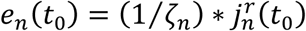 (Here, *ζ*_*n*_ is the unknown documented-to-exposed ratio for state *n*). Finally, the age-stratified compartments were initialized according to the state-wise demographic composition, i.e. *u*_*n,i*_(*t*_0_) = *f*_*n,i*_*u*_*n*_(*t*_0_). The mobility matrices *P*_*m,n*_ and passenger flux *ϕ* were calculated using the latest monthly-resolved aviation data (between September 2018 and August 2019) released by the United States Bureau of Transportation Statistics (*24*) (refer to Supplementary Materials (*25*) for details). It was assumed in our model that the interstate exchange of passengers is predominately via air traffic since volume of ground-based exchange is negligible (see Supplementary Materials (*25*)). We further assumed that the long-term international importation of individual infected with COVID-19 is minimal, under a wholesale travel restriction enforced on international passengers that arrive from countries and regions where COVID-19 is widespread (*26*-*28*). The state-wise demographic composition *f*_*n,i*_ was acquired from database of United States Census Bureau (*29*). The matrix of *T*_*i,k*_ was acquired from the work by Zagheni et al. (*15*) wherein the age-specific time-of-exposure was calculated using American Time Use Survey data (Supplementary Materials (*25*)).

The epidemiological parameters, *D*_*E*_, *D*_*I*_, and εε of COVID-19 were assumed to be 3.69 days, 3.48 days, and 1.82, respectively, per the values reported in the latest modelling study conducted on COVID-19 epidemics in China (*2*). For simplicity, we assumed here that the country-wise variability in the incubation and infectious periods of coronavirus is insignificant. The unknown state-wise parameters, *R*_0,*n*_, *η*_*n*_, and *ζ*_*n*_ were inferred in a trial-and-error manner, by fitting the model predicted 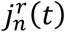 to the observational data within a calibration period. Specifically, the inference algorithm operates by iteratively guessing the values of *R*_0,*n*_, *η*_*n*_, and *ζ*_*n*_ for each state *n*, and repeats this process until attaining the optimal ***R***_***0***_, ***η***, and ***ζ*** arrays that minimize the root-mean-squared-error (RMSE) between the model predicted 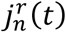 and the ground truth. The calibration period was set to the ten day period between March 19 and 28, during which a rapid surge in COVID-19 cases was observed in the US states. Such a rapid growth in epidemic size preceded an inflection point taking place at the start of April due to the manifestation of the effectiveness of country-wide social distancing (Refer to Supplementary materials (*25*) for detail of this inflection). We therefore assume that the epidemic dynamics between March 19 and 28 is representative of a baseline scenario with no intervention in effect.

## Results

### Model calibration

Figure 1 (a) and (b) shows the iterative inference results of COVID-19 epidemiological parameters. Panel (a) compares the 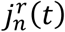 predicted by the best-fit model (line) with the observed epidemic trends in New York, California, Texas, and Washington DC (Refer to Supplementary Materials (*25*) for the complete inference results for all 52 locations). Figure 1(b) shows the combination of *η*_*n*_ and *ζ*_*n*_ that gives rise to the minimum RMSE, under a fixed *R*_0,*n*_ (taking values outlined in the respective subpanels in (a)). Fig. 1 (c) and (d) show the distribution of *R*_0,*n*_, *η*_*n*_, and *ζ*_*n*_ for all 52 locations. The nationwide median value of *R*_0_ is found to be about 4.05, with the 25^th^ and 75^th^ percentile taking values of 3.99 and 5.04, respectively. These values are consistently larger than that reported in the other modelling studies conducted on the COVID-19 epidemics in China (*30*). This greater infectiousness could be due to the absence of public awareness and effective intervention in the US at the early stage of epidemic outbreak. The nationwide median values of *η* and *ζ* are found to be about 0.7 and 0.2, respectively.

**Figure 1.**
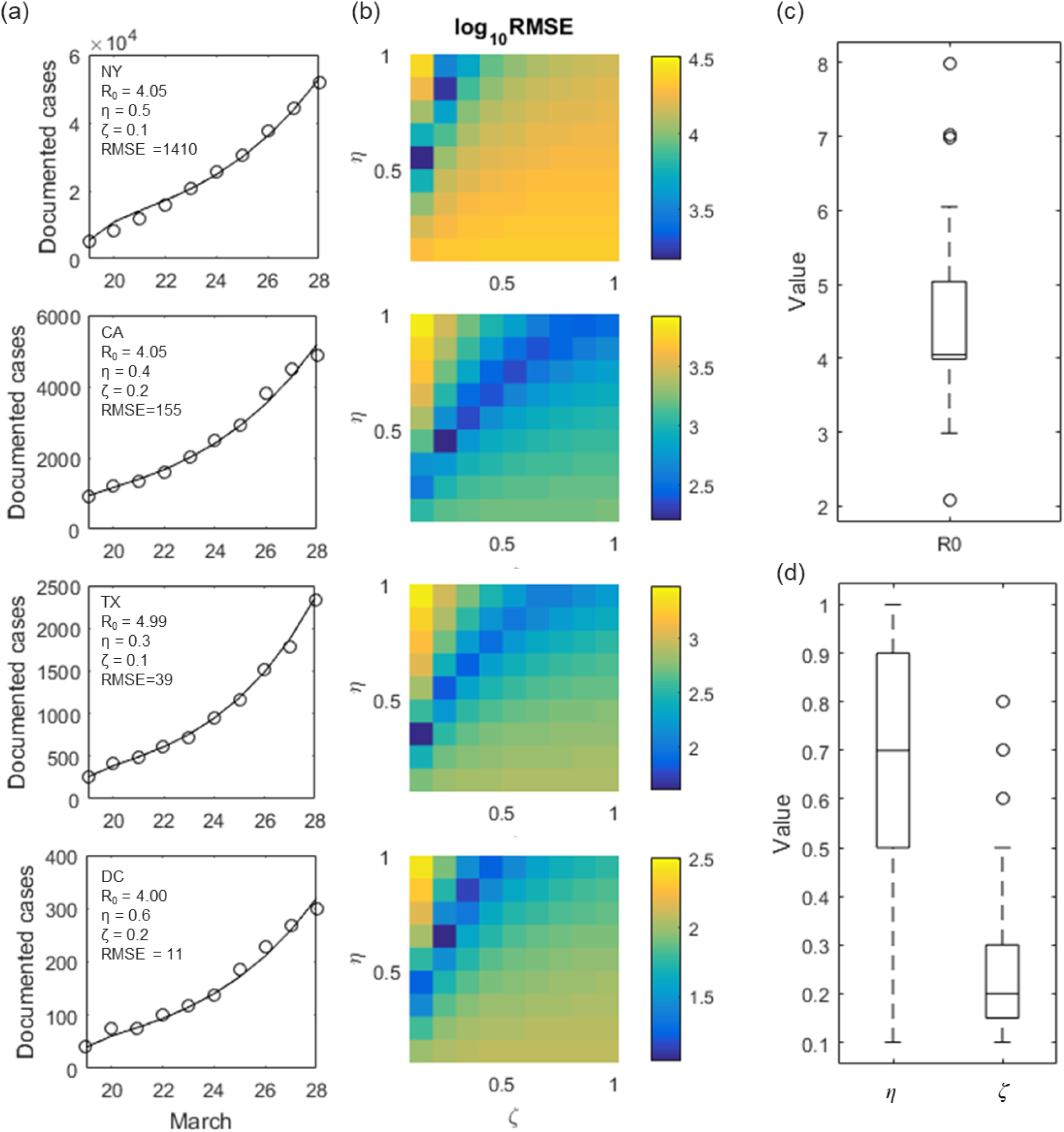
Model calibration. **(a)** Daily observation of documented active COVID-19 cases (circle) is compared with the best-fit model (line). The parameter combination leading to the best-fit is outlined in each subpanel. **(b)** Root-mean-square-error (RMSE) between observation and model prediction is plotted as a function of documentation ratio (*η*) and initial exposure ratio (*ζ*) for each corresponding state. The basic reproduction ratio (*R*_0_) is held constant here, taking the values outlined in the respective subpanels in **(a). (c)** Distribution of *R*_0_ across the 52 locations. (**d**) Distribution of *η* and *ζ* across the 52 locations. In **(c)** and **(d)**, center of the box represents state-wise median values. Edges of the box represent the 25^th^ and 75^th^ percentile. Whiskers extend to the extreme data points not considered outliers, and outliers are represented by circle symbol.

### Effectiveness of social distancing in balancing medical demand-and-supply

We estimate the demand on medical system across the country, by assuming that no effective containment intervention will take place and the epidemics will persist the trend observed between March 19 and 28. In the context of this work, we interpret the burden on medical resource using local demand-to-supply ratio for hospital beds (*y*_*H,n*_) and ICU beds (*y*_*ICU,n*_) at epidemic peaks:

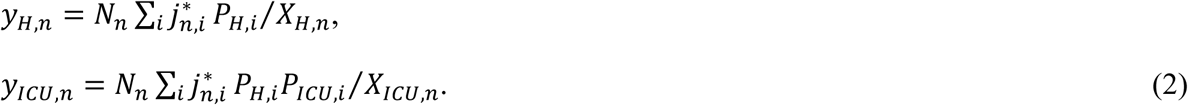

Here,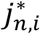 is the peak value of *j*_*n,i*_(*t*); *N*_*n*_ is the total population of state *n* (*29*); *P*_*H,i*_ and *P*_*ICU,i*_ respectively denote the age-specific rate, at which COVID-19 patients require hospitalization, and the age-specific rate, at which hospitalized cases require critical care (data tabulated in Supplementary Material (*25*), Source (*12*)); *X*_*H,n*_ and *X*_*ICU,n*_ respectively denote the state-wise numbers of available hospital beds and ICU beds (data tabulated in Supplementary Material (*25*), Source (*31*.)). Figure 2 (a) and (b) color each US state according to its corresponding *y*_*H,n*_ and *y*_*ICU,n*_ values, respectively. If the epidemic progression remains unhalted, the state-wise hospital beds and ICU beds could be overwhelmed by up to 12 and 35 times, respectively.

**Figure 2.**
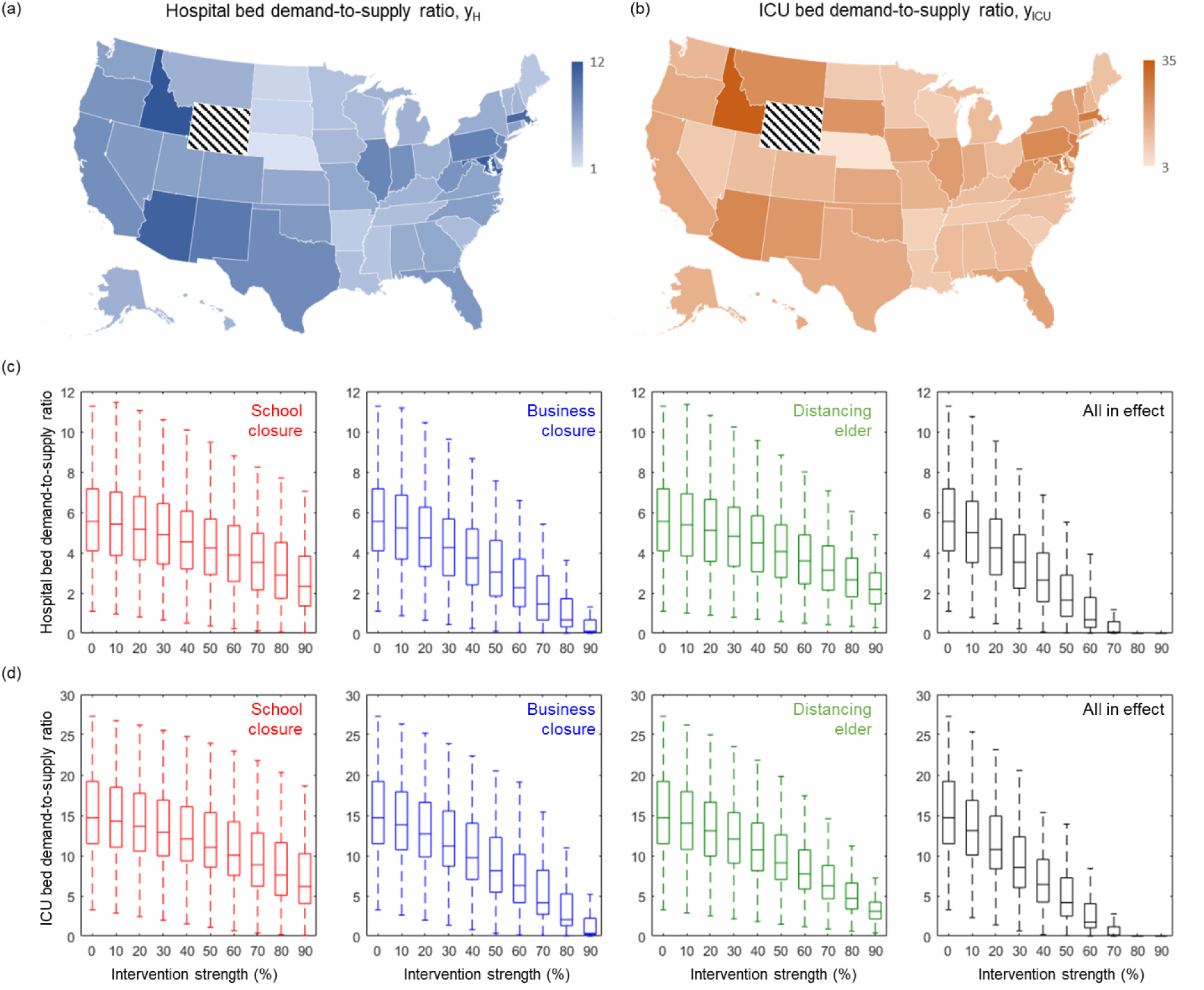
Effectiveness of various social distancing intervention in reducing hospital demand-to-supply. **(a)** and **(b)** respectively show the estimated demand-to-supply ratio of hospital beds (*y*_*H*_) and ICU beds (*y*_ICU_) at state-wise epidemic peaks. State of Wyoming (an outlier with *y*_*H*_ ≈ 16 and *y*_*ICU*_ ≈ 94) is not shown here. **(c)** Demand-to-supply ratio of hospital bed *y*_*HH*_ is plotted as a function of intervention strength (*φ*) – defined as the percent reduction in the exposure time of individuals targeted by the intervention. Center of the box represents state-wise median values of *y*_*H*_ and *y*_*ICU*_. Edges of the box represent the 25^th^ and 75^th^ percentile. Whiskers extend to the extreme data points not considered outliers. Outlier (State of Wyoming) is not shown in this figure. **(d)** Same plots for demand-to-supply ratio of ICU bed *y*_*ICU*_.

Panel (c) and (d) evaluate the effectiveness of various social distancing practices in balancing the medical demands and supply. Herein the decreases in the peak *y*_*H*_ and *y*_*ICU*_ are plotted as functions of the intervention strength (*φ*) – defined as the percent reduction in time-of-exposure *T*_*i,k*_ of the targeted age-group members. For example, school closure reduces the values of *T*_*i,k*_ elements that are associated with individuals aged 1-19 years; Business closure and distancing the elder reduce *T*_*i,k*_ for those aged 20-59 years, and those aged 60-year-and-above, respectively. Formally, this treatment is described with the following time-of-exposure matrix (*T*_*i,k*,intv_) modified per the type of social distancing practice:

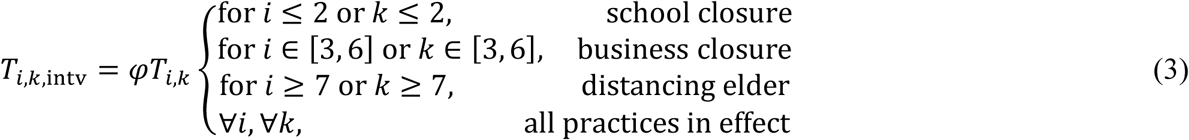

The side-by-side comparison in Fig. 3 shows that business closure is the most effective practice, possibly due to the fact that it targets at the majority of the population. At a fixed *φ* level, elder distancing achieves a better outcome than school closure. However, none of these practices could curb COVID-19 adequately, if they are implemented in a separate manner. Instead, a wholesale social distancing with all practices in effect must be taken, and it takes at least an intervention strength *φ* = 70% to reduce the medical demands to a balanced level. (hereafter, the term “social distancing” will be used to denote a wholesale distancing with *φ* = 70%).

**Figure 3.**
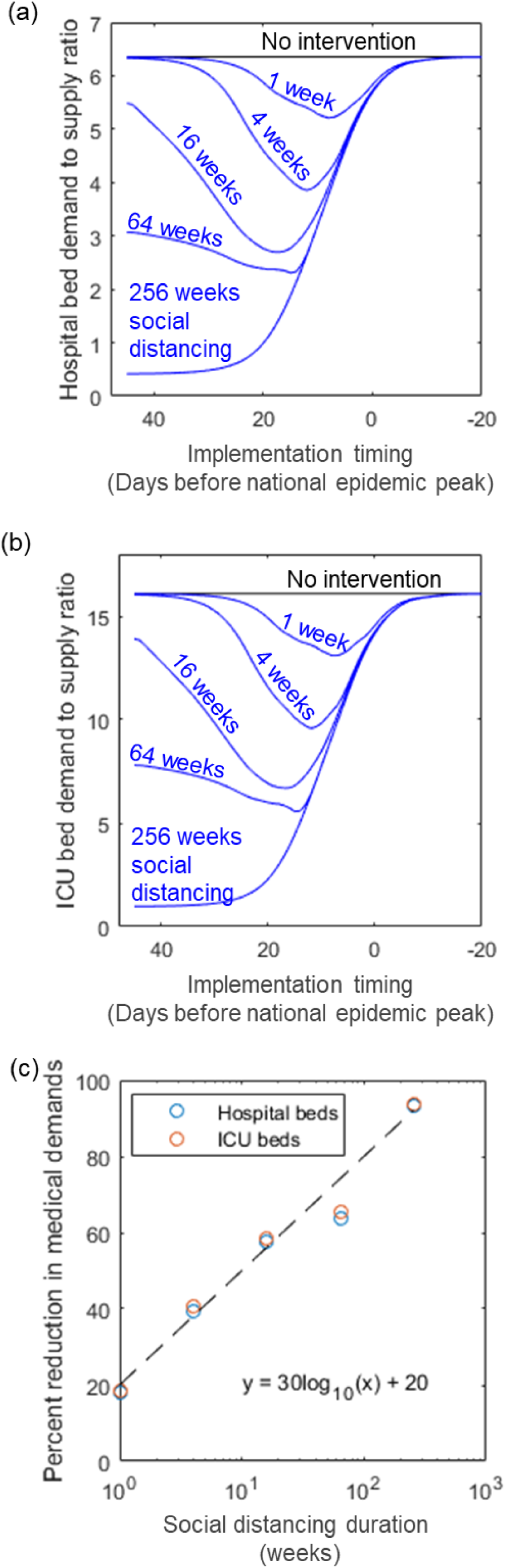
Timing and effectiveness of the finite duration social distancing. **(a)** Nationwide hospital bed demand-to-supply ratio 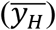 is plotted as a function of social distancing duration and implementation timing. **(b)** Same plot for nationwide ICU bed demand-to-supply ratio 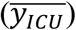. **(c)** Semi-log plot for the relationship between social distancing duration and the corresponding maximal decrease in medical demands.

### Diminishing marginal benefit of social distancing

A prolonged social distancing could have a devastating socio-economical implication that outweighs its benefits (*32,33*). If the society could only afford a finite-time social distancing, it is of utmost importance to understand *when* and for *how long* the intervention should be put into effect, so as to maximize the net benefit. Figure 3 (a) and (b) plot the effectiveness of a finite-time social distancing as functions of intervention duration and implementation timing. (Here, the nationwide demand-to-supply ratio of medical resources: 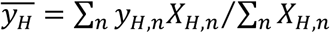 and 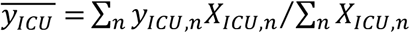 are used to benchmark intervention effectiveness). The trends in Fig. 3 (a) and (b) show that unless the intervention could last indefinitely (such as the hypothetical 256 weeks), a premature implementation could be counterproductive. Furthermore, the diminishing marginal benefits of social distancing can be inferred from these trends. For example, social distancing lasting for 1-week, 4-week, and 16-week respectively reduces the 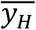 (and 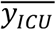) by up to 20%, 40%, and 60%: A linear decrease in medical demands is achieved at the cost of an exponential increase in social distancing duration. Such a semi-logarithm relationship between the cost and benefit of social distancing is captured in Fig. 3 (c).

We next evaluate the effectiveness of intermittent social distancing strategy – an arrangement comprising alternating phases of social distancing and normalcy that last for variable durations (*34*). Figure 4 (a) and (b) plot the medical resource demand-to-supply ratio as functions of the durations of the social distancing phase (*τ*_*D*_) and the normalcy phase (*τ*_*N*_). For example, a “weekday-like” intermittent social distancing, with a *τ*_*N*_ = 5 days and a *τ*_*D*_ = 2 days, is marked in Fig. 4 (a) and (b) with symbol “A”. Under such an arrangement, the nationwide 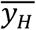 and 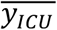 are about 5.0 and 12.6, respectively. Furthermore, if the portion of distancing phase is stretched to a level resembling a “bi-weekly” arrangement, i.e. *τ*_*N*_ = 9 days and *τ*_*D*_ = 5 days (symbol “B” in panel (a) and (b)), the nationwide 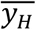 and 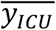 can be reduced to about 2.8 and 6.7, respectively. The trends in (a) and (b) also imply that the arrangements with identical distancing-to-normalcy ratio (*τ*_*D*_/*τ*_*N*_) tend to achieve the same outcome. This observation is further elucidated in Fig. 4 (c), wherein the reduction in medical demands is plotted as functions of the characteristic *τ*_*D*_/*τ*_*N*_ ratio. The diminishing marginal benefits of social distancing is again observed here. Up to 80% reduction in medical demands can be achieved with a *τ*_*D*_/*τ*_*N*_ ≈ 5. The benefit of further increase in *τ*_*D*_/*τ*_*N*_ is negligible.

**Figure 4.**
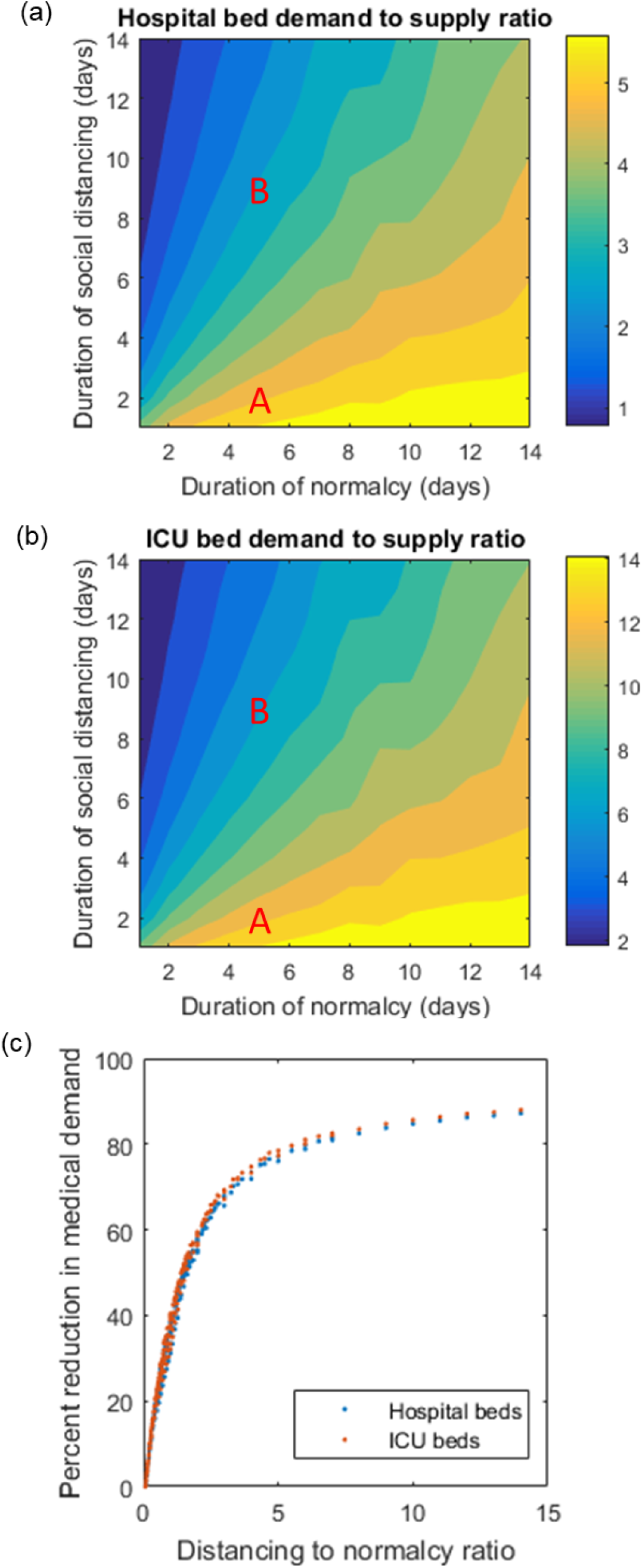
Effectiveness of intermittent social distancing with variable distancing phase and normalcy phase duration. **(a)** Nationwide hospital bed demand-to-supply ratio 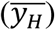 is plotted as a function of social distancing phase duration (*τ*_*D*_) and normalcy phase duration (*τ*_*N*_). **(b)** Same plot for nationwide ICU bed demand-to-supply rati 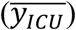. Symbol “A” marks a “weekday-like” arrangement comprised by intermittent five-day normalcy and two-day social distancing. Symbol “B” marks a “bi-weekly working arrangement” comprised by intermittent five-day normalcy and nine-day social distancing. **(c)** Percent reduction in medical demands is plotted as a function of distancing to normalcy ratio, *τ*_*D*_/*τ*_*N*_.

## Discussion

In this work, we provide a comprehensive systematic analysis of the effectiveness of social distancing in alleviating the burden of COVID-19 on nationwide medical resources. Our baseline scenario represents a continuation of the epidemic dynamic between March 19 and 28, during which a rapid surge of COVID-19 cases is observed in the United States and the effect of social distancing is yet to manifest. Our finding suggests that under such a baseline condition, the state-wise hospital and ICU beds could be overwhelmed by up to 12 and 35 times, respectively. A wholesale social distancing could balance the medical demand-to-supply at epidemic peak provided a 70% percent reduction in the time-of-exposure of the population within all age-groups is achieved. Marginal benefits of social distancing wane over time, characterized by a linear decrease in medical demand against exponentially increasing social distancing duration. We therefore suggest an intermittent social-to-no-distancing arrangement, which could reduce the medical demands by up to 80% provided the duration of social distancing is less than or equal to five times of the recurring normalcy period. Findings from this study may also apply to other regions of Europe, as well as Asia, where social distancing measures have been in effect to slow the epidemic spread.

## Data Availability

All Processed data are made available in the Supplementary Materials of the manuscript and online via the website: https://eece.wustl.edu/chakrabarty-group/covid/

